# Coronavirus disease-19: Summary of 2,370 Contact Investigations of the First 30 Cases in the Republic of Korea

**DOI:** 10.1101/2020.03.15.20036350

**Authors:** COVID-19 National Emergency Response Center, Epidemiology and Case Management Team, Korea Centers for Disease Control and Prevention

## Abstract

Between January 24 and March 10, of a total of 2,370 individuals who had contacted the first 30 cases of COVID-19, 13 were found to have COVID-19, resulting secondary attack rate of 0.55% (95% CI 0.31 – 0.96). Of 119 household contacts, 9 had infections resulting secondary attack rate of 7.56 (95% CI 3.73 – 14.26).

## Introduction

Since the importation of coronavirus disease 2019 (COVID-19) in the Republic of Korea, the first 30 confirmed cases were reported as of February 17th, 2020. The epidemiological and clinical characteristics of 28 cases of those were analyzed in previous report (1).

Tracing of contacts of cases is essential for containing COVID-19 within the community. In Korea, an established system exists using public health centers to conduct epidemiological investigation and early quarantine/isolation of suspected case, thus interrupting the line of transmission. This approach has been particularly successful in containing the COVID-19 in early phase of outbreak.

Here, we describe the summary of 2,370 contact investigations of the first 30 cases of coronavirus disease-19 (COVID-19) in Korea. The study includes a secondary attack rates among different age groups and mode of transmission.

## Materials and Methods

Demographic, epidemiological, and early clinical information were retrieved from COVID-19 reporting and surveillance data from Korea Centers for Disease Control and Prevention (KCDC). As in previous report, patient age was provided on the date of diagnosis, and key indicators were identified by an epidemiological investigator who participated in the field investigation and the epidemiological investigation team (1). It must be noted that the data presented in this study may change depending on the results of further epidemiological investigation.

Initial working definition for ‘close contact (or high risk exposure)’ was defined as being within approximately 2 meters of a COVID-19 case; and definition for ‘daily contact (or low risk exposure)’ was defined as having proximity of a person with confirmed COVID-19 case, without having had close contact. The classification was then repealed and was integrated into ‘contact (regardless of level of exposure)’. All asymptomatic contacts were mandated to stay self-quarantine for 14 days and were put under active surveillance by public health workers who called twice a day to check presence of fever or respiratory symptoms. Smartphone-based ‘self-assessment app’ was introduced as an additional tool to track the symptoms.

## Result

Between January 24 and March 10, of a total of 2,370 individuals who had contacted the first 30 cases of COVID-19, 12 were found to have COVID-19, resulting secondary attack rate of 0.55% (95% CI 0.31 – 0.96) (Table 1). Male had higher attack rate (0.75, 95% CI 0.38 – 1.47) than female (0.38, 95% CI 0.16 – 0.89). Of 119 household contacts, 9 had infections resulting secondary attack rate of 7.56% (95% CI 3.73 – 14.26) (Table 1). Number of traced individuals ranged from 15 to 649 persons and mean monitored counts ranged from 5.7 to 31.3 times (Table 2).

**Table 1.** Demography of traced individuals and secondary attack rates, coronavirus diseases-19, Republic of Korea

| Variables | Traced<br>*N* | Infected<br>*N* | Secondary attack rate |  |
| --- | --- | --- | --- | --- |
|  |  |  | % | (95% CI) |
| Sex |  |  |  |  |
| Female | 1,303 | 5 | 0.38 | (0.16-0.89) |
| Male | 1,067 | 8 | 0.75 | (0.38-1.47) |
| Age group (years) |  |  |  |  |
| 0-9 | 88 | 0 | - | - |
| 10-19 | 67 | 1 | 1.49 | (0.08-9.13) |
| 20-29 | 486 | 3 | 0.62 | (0.21-1.80) |
| 30-39 | 477 | 0 | - | - |
| 40-49 | 439 | 3 | 0.68 | (0.23-1.99) |
| 50-59 | 388 | 3 | 0.77 | (0.26-2.25) |
| 60-69 | 275 | 2 | 0.73 | (0.20-2.61) |
| 70-79 | 91 | 1 | 1.10 | (0.19-5.96) |
| ≥80 | 45 | 0 | - | - |
| Unknown | 20 | 0 | - | - |
| Mode of contact |  |  |  |  |
| Household contacts* | 119 | 9 | 7.56 | (3.73-14.26) |
| Others | 2,151 | 4 | 0.19 | (0.07-0.48) |
| Total | 2,370 | 13 | 0.55 | (0.31-0.96) |

\*Provisional data, as of March 13 2020

**Table 2.** Number of traced individuals and secondary transmissions by cases, coronavirus disease-19, Republic of Korea

| Case No. | Traced Individuals N | Monitored days, mean (/person) | Infected Individuals N | Transmission Description |
| --- | --- | --- | --- | --- |
| 3 | 95 | 16.7 | 1 | Transmitted to case #6 (meal) |
| 5 | 35 | 17.0 | 1 | Transmitted to case #9 (meal) |
| 6 | 23 | 16.8 | 3 | Household transmission to cases #10, #11<br>Transmitted to case #21 (church) |
| 12 | 649 | 9.1 | 1 | Household transmission to case #14 |
| 15 | 15 | 31.3 | 2 | Transmitted to case #20 (meal) |
| 16 | 452 | 16.0 | 2 | Household transmission to cases #18, #22 |
| 20 | 2 | 12.0 | 1 | Household transmission to case #32 |
| 27 | 40 | 5.7 | 2 | Household transmission to cases #25, #26 |
| 29 | 111 | 19.9 | 1 | Household transmission to case #30 |

\*Provisional data, as of March 13 2020

## Discussion

Tracing of 2,370 contacts of the first 30 COVID-19 cases in Korea indicate that the risk of symptomatic transmission among contacts was low at 0.55% (95% CI 0.31 – 0.96). However, the findings also suggested that the transmission of COVID-19 was significant among household contacts, which is in line with other reports. In the earlier reports, familial clusters had been reported and was there were question on whether household transmission could be a major driver in spread of outbreak in the community (2, 3). Of the first 262 COVID-19 cases in Beijing, China, 133 (50.8%) were family cluster cases (4). In the US, an active symptom monitoring was performed for 445 close contacts of the 12 cases with travel-related COVID-19, resulting a symptomatic secondary attack rate of 0.45% (95% CI, 0.12 – 1.6) among all contacts, and 10.5% (95% CI, 2.9 – 31.4) among household members (5).

This study suggest that contact tracing was critical for containing the COVID-19 outbreak in the early phase in Korea. Contact tracing rely on other concurrent aspects of the COVID-19 containment strategies such as investigating, classifying, tracking, and managing contacts by identifying the patient’s route. A mathematical model suggested highly effective contact tracing and case isolation was enough to control a new outbreak of COVID-19 in most scenarios (6). In Korea, various measures such as tracking the history of clinic visits, GPS of the cell phones, credit card transaction log, and CCTV had been utilized to complete the contact tracing of COVID-19 cases (7).

There are certain limitations that should be considered. First, this is a summary of the first 30 cases of COVID-19 in Korea, when containing of the cases and contacts was the mainstay of control strategy. Following the increase of cases after case number 31, the strategy has shifted from containment to mitigation, which is now applied in many parts of the world. Second, some potential risk factors were not assessed and would merit further study, such as the characteristics of the household and other transmission routes.

However, the result underscores the infectious nature of COVID-19 in the household and identify the main driver that facilitate secondary transmission in the community. This summary identifies the overall risk of symptomatic transmission of COVID-19 in the setting of early importation phase of the disease. Implementation of basic infection control practices, such as isolation of ill family members and enhancement of hand hygiene measures, may lead to a reduction in transmission. Future analyses should attempt to incorporate screening of COVID-19 and serology in symptomatic and asymptomatic contacts.

## Data Availability

Epidemiologic investigation files from Korea CDC

## Acknowledgments

We thank the relevant ministries, including the Ministry of Interior and Safety, Si/Do and Si/Gun/Gu, medical staffs in health centers, and medical facilities for their efforts in responding to COVID-19 outbreak.

## Conflicts of Interest

The authors declare no competing financial interests.

## Notes

### Competing Interest Statement

The authors have declared no competing interest.

### Funding Statement

None exist

